# Transferrin receptor-based circulating tumor cell enrichment provides a snapshot of the molecular landscape of solid tumors and correlates with clinical outcomes

**DOI:** 10.1101/2024.06.16.24309003

**Authors:** Giuseppe Galletti, Ahmed Halima, Ada Gjyrezi, Jiaren Zhang, Bob Zimmerman, Daniel Worroll, Galatea Kallergi, Rohan Barreja, Allyson Ocean, Ashish Saxena, Timothy E. McGraw, David M. Nanus, Olivier Elemento, Nasser K. Altorki, Scott T. Tagawa, Paraskevi Giannakakou

## Abstract

Circulating tumor cells (CTCs) captured from the bloodstream of patients with solid tumors have the potential to accelerate precision oncology by providing insight into tumor biology, disease progression and response to treatment. However, their potential is hampered by the lack of standardized CTC enrichment platforms across tumor types. EpCAM-based CTC enrichment, the most commonly used platform, is limited by EpCAM downregulation during metastasis and the low EpCAM expression in certain tumor types, including the highly prevalent and lethal NSCLC. In this study we demonstrate that Transferrin Receptor (TfR) is a selective, efficient biomarker for CTC identification and capture in patients with prostate, pancreatic and NSCLC. TfR identifies significantly higher CTC counts than EpCAM, and TfR^+^-CTC enumeration correlates with disease progression in metastatic prostate and pancreatic cancers, and overall survival and osimetrinib-resistance in non-small cell lung cancer (NSCLC). Profiling of TfR^+^-CTCs provides a snapshot of the molecular landscape of each respective tumor type and identifies potential mechanisms underlying treatment response to EGFR TKi and immune checkpoint inhibitors in NSCLC.

**One sentence summary:** Transferrin Receptor identifies circulating tumor cells in solid tumors

## Introduction

Circulating tumor cells (CTCs) have emerged as a validated source of tumor cells which are obtained non-invasively with a simple blood draw and allow real-time monitoring of disease progression and therapeutic response (*1–4*). Importantly, CTC analysis is not subject to the bias of biopsy site while reflecting tumor cells shed from both primary tumor and metastatic sites. Thus, CTC analysis provides a more comprehensive picture of the disease burden and heterogeneity.

Currently, the main clinical application of CTCs is limited to enumeration used as a prognostic biomarker approved in only three tumor types (breast, prostate and colorectal) (*5–7*). As CTCs represent tumor cells with intact DNA, RNA and protein content, unlike the fragmented cfDNA, they can provide comprehensive information about the tumors they disseminate from, which can be used in precision oncology. For example, we and others have demonstrated that transcriptomic CTC profiling can identify gene signatures predictive of clinical response to standard of care treatments (*8, 9*). However, such a treatment customization approach has not been clinically adopted due to the lack of a uniform CTC isolation platform. Currently, CTC positive selection relies largely on the use of EpCAM, the epithelial specific cell surface protein. However, EpCAM-based CTC detection is limited by EpCAM’s downregulation during epithelial-to-mesenchymal transition (EMT), a process that precedes metastatic dissemination (*10*), as well as by its low expression in select tumor types. Non-small cell lung cancer (NSCLC), a highly prevalent disease and the leading cause of cancer death in the US, is such a tumor type where EpCAM has no clinical use (*11, 12*). Thus, the identification of a novel cell surface protein overexpressed in tumors as a pan-cancer CTC identifier remains a clinically unmet need.

To address this gap, we investigated the role of transferrin receptor (TfR), a plasma membrane protein that mediates iron uptake and is overexpressed in solid tumors (*13, 14*), as an alternative to the EpCAM CTC identifier. The selective overexpression of TfR in cancer cells, the clinical association of TfR with advanced disease stage and worse prognosis in solid tumors, and its plasma membrane localization (*49, 50*) have made TfR an attractive target as a diagnostic and therapeutic resource (*51–53*). These characteristics also make TfR also an ideal candidate for the identification and isolation of CTCs. We investigated TfR-based CTC enrichment in patients with prostate and pancreatic cancer, two tumor types where EpCAM-based CTC enumerations is approved or feasible, respectively, as well as in NSCLC, where EpCAM-based CTC isolation has no clinical use. We show that in patients with prostate and pancreatic cancer, TfR identifies significantly higher counts of CTCs which are molecularly distinct from their EpCAM^+^ counterparts. Remarkably, we also show that TfR identifies CTCs in both early-stage and metastatic NSCLC, and that their enumeration and molecular characterization correlates with clinical outcomes and has the potential to inform therapeutic strategies.

## Results

### TfR expression is more prevalent than EpCAM across tumor types, is not affected by EMT, and correlates with disease stage in prostate cancer

To determine the suitability of TfR as a biomarker for CTC enrichment in solid tumors we first characterized TfR performance in cell lines in comparison to EpCAM. Using a prostate cancer model of inducible EMT (*27*) we showed that TfR expression was not affected upon induction while EpCAM expression was lost (**Supplementary Figure 1A-B**). In addition, we tested TfR and EpCAM expression and plasma membrane association in 22 cell lines from diverse solid tumors. We found that TfR was expressed in the majority of cell lines (21/22 or 96%) while EpCAM was present in 11/22 (50%) (**Supplementary Figure 2 and Supplementary Table 1**). To test the suitability of TfR for CTC capture we examined the dynamic range of TfR staining in healthy donor peripheral blood spiked with cancer cells, as a surrogate for CTCs. Flow cytometry and immunofluorescence analyses showed that TfR expression was not detected in the CD45^+^ leukocytes (**Supplementary Figure 3A-C**) while it was highly expressed in epithelial cancer cells. This result is consistent with a recent report showing that TfR is a cancer cell affinity target in blood samples with no staining interference from other blood cells (*28*). Next, we investigated TfR RNA expression by mining publicly available large datasets. These datasets were the TCGA dataset (*29*), consisting of 505 primary prostate cancer samples, the SU2C dataset (*30*), and our internal dataset consisting of 91 metastatic castration-resistant prostate cancer (mCRPC) patient samples. Our analysis showed that TfR expression increased with disease stage from localized to metastatic CRPC (22% vs 68% above the mean) (**Figure 1A**), while EpCAM expression remained largely unchanged (46% vs 40% above the mean) (**Figure 1B**). A similar pattern was obtained when we analyzed our internal cohort of 91 patient samples, including 39 primary PC, 32 CRPC and 20 neuroendocrine PC (NEPC) samples (*21*) (**Figure 1C**). TfR expression was significantly increased with disease stage, while EPCAM expression did not change.

**Figure 1.**
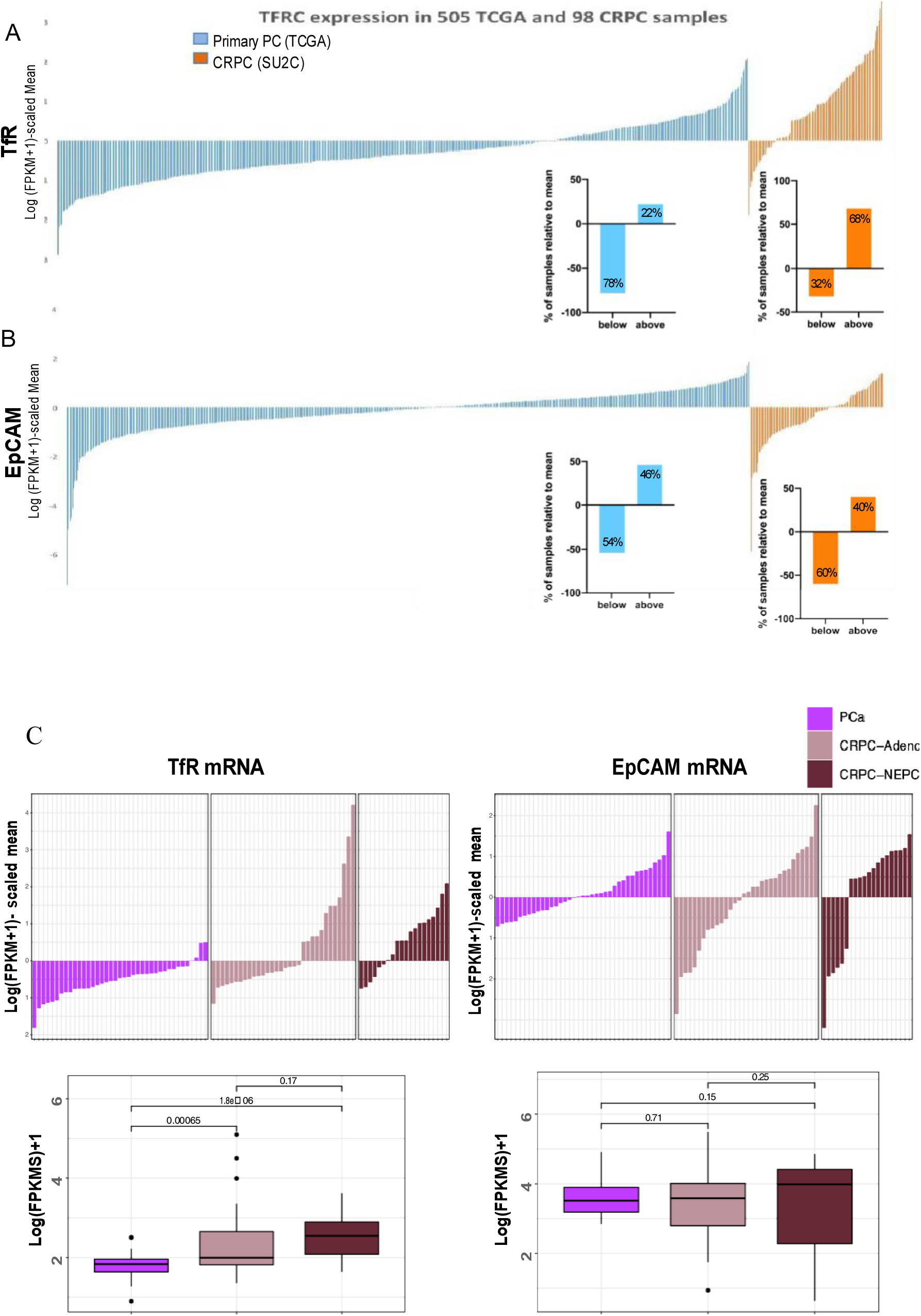
Prevalence and distribution of TfR and EpCAM expression in large clinical datasets from patients with localized or metastatic prostate cancer. (**A**) Expression of TfR (**A**) or EpCAM (**B**) was determined in primary prostate cancer samples (TCGA, blue, n=505) and metastatic CRPC (SU2C, orange, n=98) by mining RNA-seq data. The waterfall plots display the expression of the two transcripts relative to the mean. The mean expression of each transcript (scaled mean) was calculated across the two cohorts of samples and set as 0. The enclosed bar graphs show the percent distribution of each transcript relative to the scaled mean. TfR mRNA expression levels (top panel) increase from 22% above the mean in primary prostate cancer to 68% in mCRPC; on the contrary, EpCAM mRNA expression levels did not vary. (**C**) Expression of EpCAM and TfR was determined in patients with localized PC (purple, n=39), metastatic CRPC (light pink, n=32) or NEPC (burgundy, n=20) by RNA-seq. The waterfall plots were built as described for A and B. The boxplots represent the normalized expression of the transcripts in each patient cohort. Wilcoxon statistical test was used. TfR expression significantly increases going from localized PC to metastatic CRPC and the most advanced and clinically aggressive NEPC. EpCAM levels remain constant during disease progression.

### TfR-based CTC enumeration and molecular characterization correlates with disease status in patients with mCRPC

In a prospective clinical study of patients with mCRPC (n=32) (Table S2, clinical characteristics) we sought to compare head-to-head TfR-*vs* EpCAM-based CTC enumeration in the same blood draw. CTCs were enriched via negative depletion of CD45^+^-cells and labeled live with antibodies against the cell surface proteins, TfR, EpCAM and CD45 (**Supplementary Figure 4A**). In a different cohort of mCRPC, we recently used the same enrichment protocol and established the prostate cancer origin of CTCs using a stringent transcriptomic pipeline (*9*). In the current cohort of 32 mCRPC patients, CTCs were enumerated based on their respective immunophenotypes as TfR^+^/EpCAM^-^/CD45^-^ (hereafter TfR^+^), EpCAM^+^/TfR^-^/CD45^-^ (hereafter EpCAM^+^) and TfR^+^/EpCAM^+^/CD45^-^ (double positive). Our analysis showed that TfR^+^-CTCs (mean= 593 CTCs; median= 370 CTCs, range= 0-3299) were significantly more abundant than EpCAM^+^-CTCs (mean= 36 CTCs; median= 21 CTCs, range= 5-240; p <0.0001) (**Figure 2A).** Double positive TfR^+^/EpCAM^+^ CTCs were also detected in several samples, but they were the least abundant of the three CTC subpopulations (mean= 49 CTCs, median= 10 CTCs, range: 0-491). Direct comparison of matching TfR^+^ *vs* EpCAM^+^-CTCs in each individual patient, revealed that TfR^+^-CTCs were the predominant CTC subpopulation with TfR^+^-CTCs counts being up to 550-fold (mean 43-fold) higher than EpCAM^+^-CTCs (**Figure 2B and Supplementary Table 3**). In addition, there was no correlation between TfR^+^ and EpCAM^+^ CTC counts, and both appeared to be independent of disease burden (**Figure 2C**). Notably, patients with active disease at the time of blood draw had the highest TfR^+^-CTC counts, a pattern that was not observed with EpCAM^+^-CTC counts (**Figure 2D and E**). These data suggest that TfR^+^-CTCs positively correlate with disease progression. The androgen receptor (AR) splice variants AR-V7 and AR-v567es (*31*) have been associated with disease progression and resistance to treatment with AR signaling inhibitors or taxanes (*4, 22, 32*). Thus, we examined AR variant expression in TfR^+^-versus EpCAM^+^-CTCs. We isolated single CTCs (sCTCs) based on their respective immunophenotypes, as TfR^+^ or EpCAM^+^ and subjected them to ddPCR to quantify AR, AR-V7 and ARv567 mRNA expression, as we recently described (*31*). We analyzed a total of 165 single CTCs (TfR+, n=102 or EpCAM^+^, n=63) isolated from 3 different patients with mCRPC. Surprisingly, the EpCAM^+^-CTCs did not express AR-V7 (0/21 sCTCs) or ARv567 (0/21 sCTCs) while TfR^+^-CTCs expressed both variants with AR-V7 detected in 7/34 sCTCs (21%) and AR-v567es in 6/34 sCTCs (18%). As expected, AR-fl was detected in both CTC subpopulations at the same rate (24%) (**Figure 2F**).

**Figure 2.**
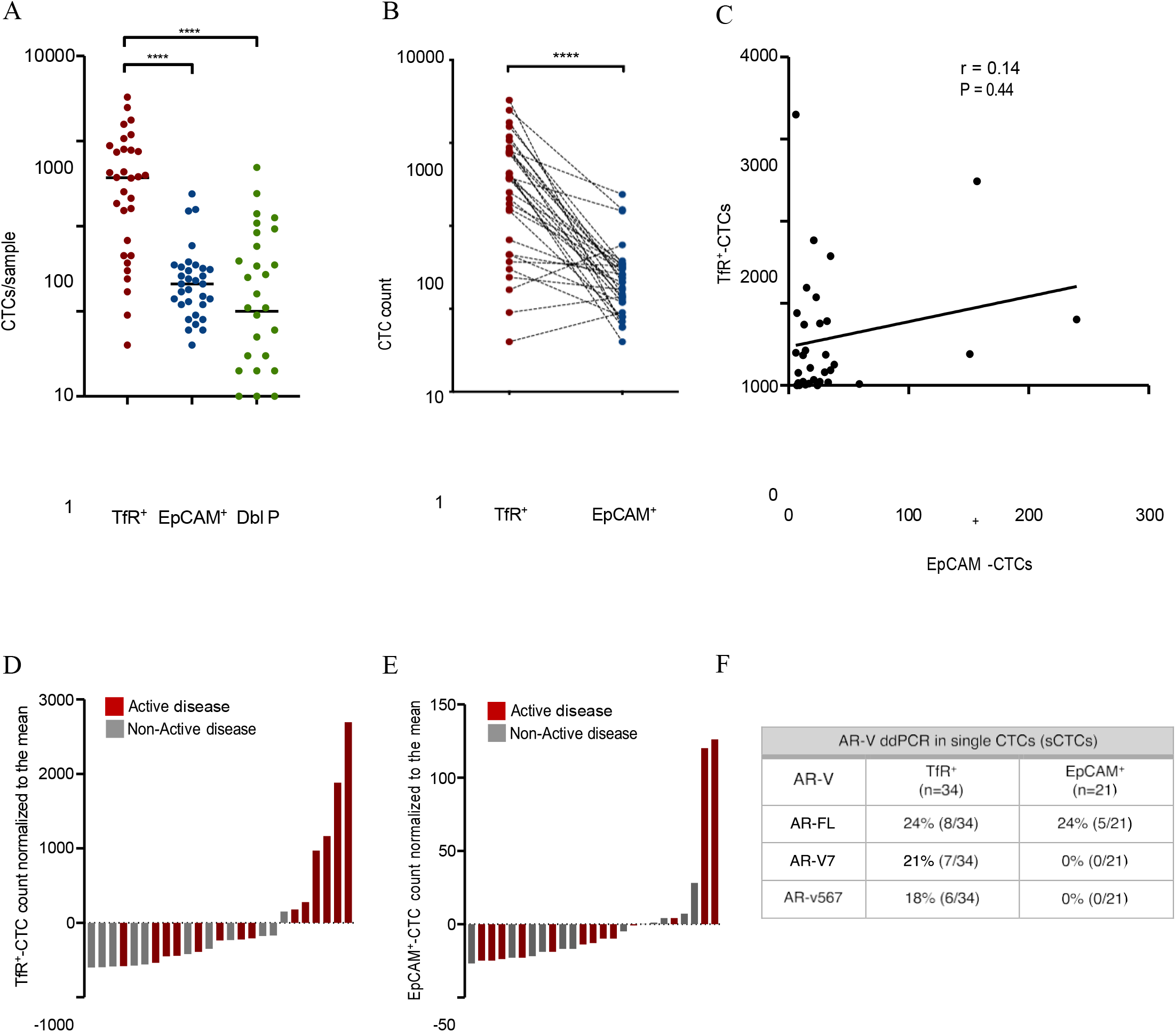
Enumeration of different TfR^+^-versus EpCAM^+^-CTC subpopulations isolated in patients with mCRPC. CTCs were enriched by negative depletion of CD45^+^-cells using peripheral blood samples from patients with mCRPC (n=32). The pool of enriched CTCs was labeled live for the cell surface antigens TfR, EpCAM and CD45, to exclude contaminating leukocytes from the analysis. CTCs were identified based on size and shape (bright field) and assigned into three immunophenotypic categories as TfR^+^/EpCAM^-^ (hereafter TfR^+^), EpCAM^+^/TfR^-^ (hereafter EpCAM^+^) and TfR^+^/EpCAM^+^ (Double positive). CTCs were enumerated using the automated microscopy-based single cell selection platform CellCelector (**A and B**) Dot plots show the CTC counts per patient for each subpopulation, and paired TfR^+^-CTCs and EpcAM^+^-CTC counts for each patient; Mann Whitney and Wilcoxon statistical tests were used. ****=p<0.0001. (**C**) Linear correlation between TfR^+^-CTCs and EpCAM^+^-CTCs in patients with mCRPC. (**D and E**) Waterfall plots show the distribution of TfR^+^- and EpCAM^+^-CTC counts from 25 patients with mCRPC for whom disease status at the time of blood draw was available. The mean number of CTC counts obtained with TfR^+^ (range: 4-3299; mean: 604) or with EpcAM^+^ (range: 4-157; mean: 31) was calculated for each CTC subpopulation and set at 0. Patients with higher than the mean CTC counts are shown as positive values and below as negative. Disease status at the time of blood draw is color coded as red (active disease) or grey (inactive). Note different CTC-count scale (Y axis) for TfR^+^-*versus* EpCAM^+^-CTCs. (**F**) Single TfR^+^-CTCs (n=102) or EpCAM^+^-CTCs (n=63) were isolated from 3 patients with mCRPC using the microscopy-based CellCelector micromanipulator and processed by ddPCR for expression of AR-FL, AR-V7 or AR-v567 transcripts (one cell/transcript) as previously described (Gjyrezi A et al. Commun Biol 2021). Equal groups of single TfR^+^-CTCs (n=34) or single EPCAM^+^-CTCs (n=21) were analyzed by ddPCR for each of the three transcripts.

Taken together these results demonstrate that, compared to EpCAM, TfR^+^-CTCs represent the predominant CTC subpopulation, and that high TfR^+^-CTCs correlate with active disease status. Importantly, TfR^+^-CTCs are enriched in AR-V7 expression, which is a clinically actionable biomarker.

### TfR identifies CTCs in early-stage and metastatic NSCLC

As mentioned earlier, EpCAM^+^-based CTC identification has not been clinically informative in NSCLC (*12*). In fact, there is no CTC enrichment platform that consistently and reliably detects CTCs in NSCLC patients. With the ever-growing number of targeted treatments for this disease, the absence of CTC-based molecular analyses is a missed opportunity for precision oncology. Thus, identification of new specific biomarkers to capture viable CTCs is an unmet clinical need in thoracic oncology.

To determine whether TfR could identify CTCs in this disease, we first analyzed blood samples from patients with resectable, early-stage disease (n=21). CTCs enriched by negative depletion were stained for TfR and cytokeratin (CK), to confirm the epithelial cell origin, as well as CD45 to exclude any contaminating leucocytes. To nominate putative CTCs, we enumerated TfR^+^/CK^+^/DAPI^+^/CD45^-^ cells (**Figure 3A-B and Supplementary Table 4**). Our results showed that all 21 patients (100%) had TfR^+^-CTCs ranging from 2-43 CTCs per patient (mean: 15; median:14). Nearly all TfR^+^ cells were also CK^+^, confirming their epithelial origin. To further confirm lung origin, we co-stained a subset of samples (n=12) with TTF1, a marker routinely used in pathology to confirm lung adenocarcinoma. Our results revealed a significant positive correlation between TTF1^+^ and TfR^+^-CTCs counts (r=0.98; p<0.0001) (**Figure 3C**). Furthermore, these results demonstrate that the plasma membrane-localized TfR can be used to identify and capture live CTCs in patients with early stage NSCLC, as opposed to CK and TTF1 which are both intracellular proteins, detectable following fixation and immunostaining and hence, not suitable for live CTC enrichment. These results were confirmed by immunostaining of 11 tumor organoids derived from patients with early-stage NSCLC adenocarcinoma, where we observed co-expression of TfR, CK and TTF1 (**Figure 3D**), as expected for lung epithelial cells, and further solidifying TfR as a *bona-fide* marker for NSCLC CTCs.

**Figure 3.**
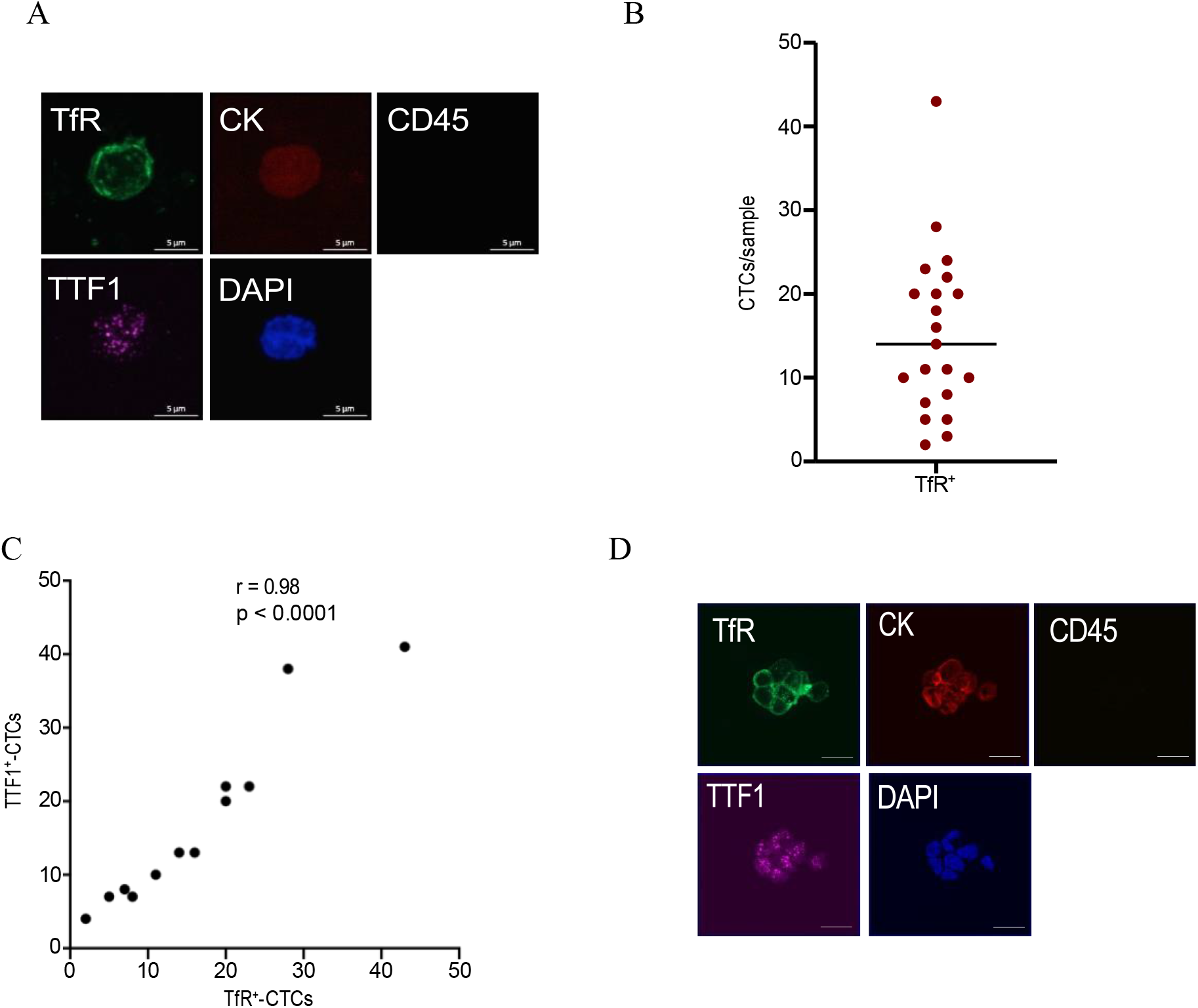
Enumeration of immunophenotypic CTC subpopulations isolated from patients with early-stage NSCLC. Blood samples were collected from chemo-naive patients with early-stage NSCLC (n=21) before surgical resection. (**A**) After CD45^+^ depletion the enriched CTC fraction was fixed and immunostained for TfR, cytokeratin (CK), TTF1 (marker of lung origin), CD45 and DAPI. (**B**) CTCs were identified based on the standard definition as CD45-/DAPI+/CK+ and the number of TfR^+^-CTCs is shown for each patient. (**C**) TTF1 staining was performed in a subset of patients (n=12) and correlated with TfR^+^ expression in single CTCs for each patient. (**D**) Organoids established using tumors resected from patients with early-stage NSCLC (n=11), were processed with the same immunostaining protocol as in A and imaged by confocal microscopy. Representative images from 1/11 organoids are displayed. Similar staining pattern was observed across all 11 NSCLC organoids. Note the similar co-expression pattern with patient-derived CTCs. Scale bar: 10 μm.

Next, we expanded our analysis to a cohort of patients with treatment-naïve metastatic NSCLC (mNSCLC) (n=35, Greek cohort) where CTCs were enriched based on size by filtration (ISET) (**Figure 4A**). CTCs were fixed and stained with antibodies against TfR, CK and CD45, as above (**Supplementary Figure 5A**). Remarkably, TfR^+^-CTCs were the most prevalent and abundant CTC subpopulation, identified in 31/35 patient samples (88%) (CTC count/sample, mean: 21; median: 6, range: 0-100) (**Figure 4B**). Using the median CTC count of 6 we dichotomized this patient cohort into TfR^+^ high (CTC ≥ 6) or TfR^+^ low (CTC < 6). We found that patients with TfR^+^ high scores had significantly shorter overall survival (3.6 vs 6.6 months, p=0.048) (**Figure 4C**). No correlation with OS was found when we used CK+ as a positive CTC identifier (p=0.426). Taken together these data demonstrate that TfR can identify lung cancer cells in the peripheral blood of patients with early and mNSCLC. Further, in the metastatic setting, TfR^+^-CTC enumeration may be prognostic.

**Figure 4.**
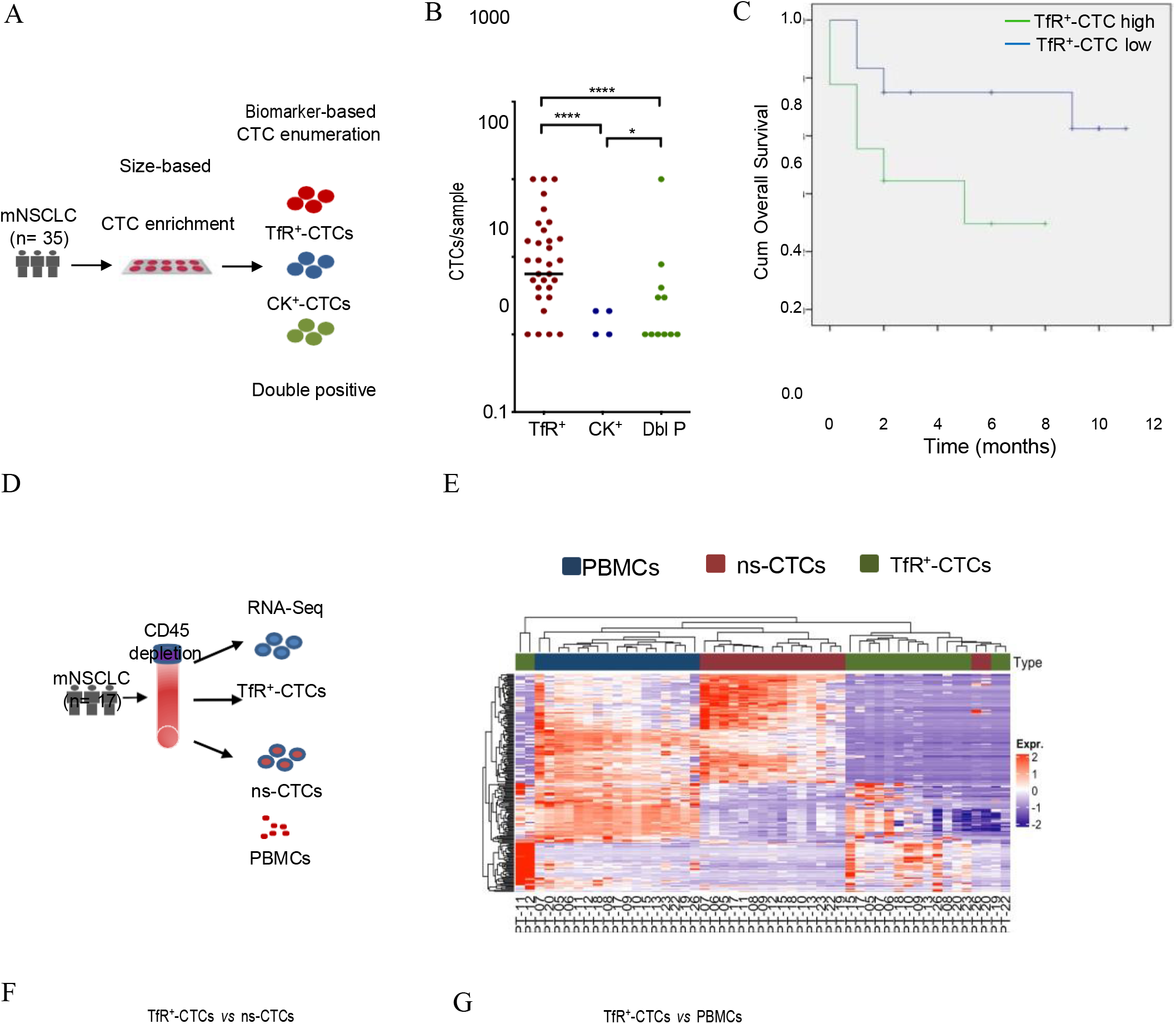
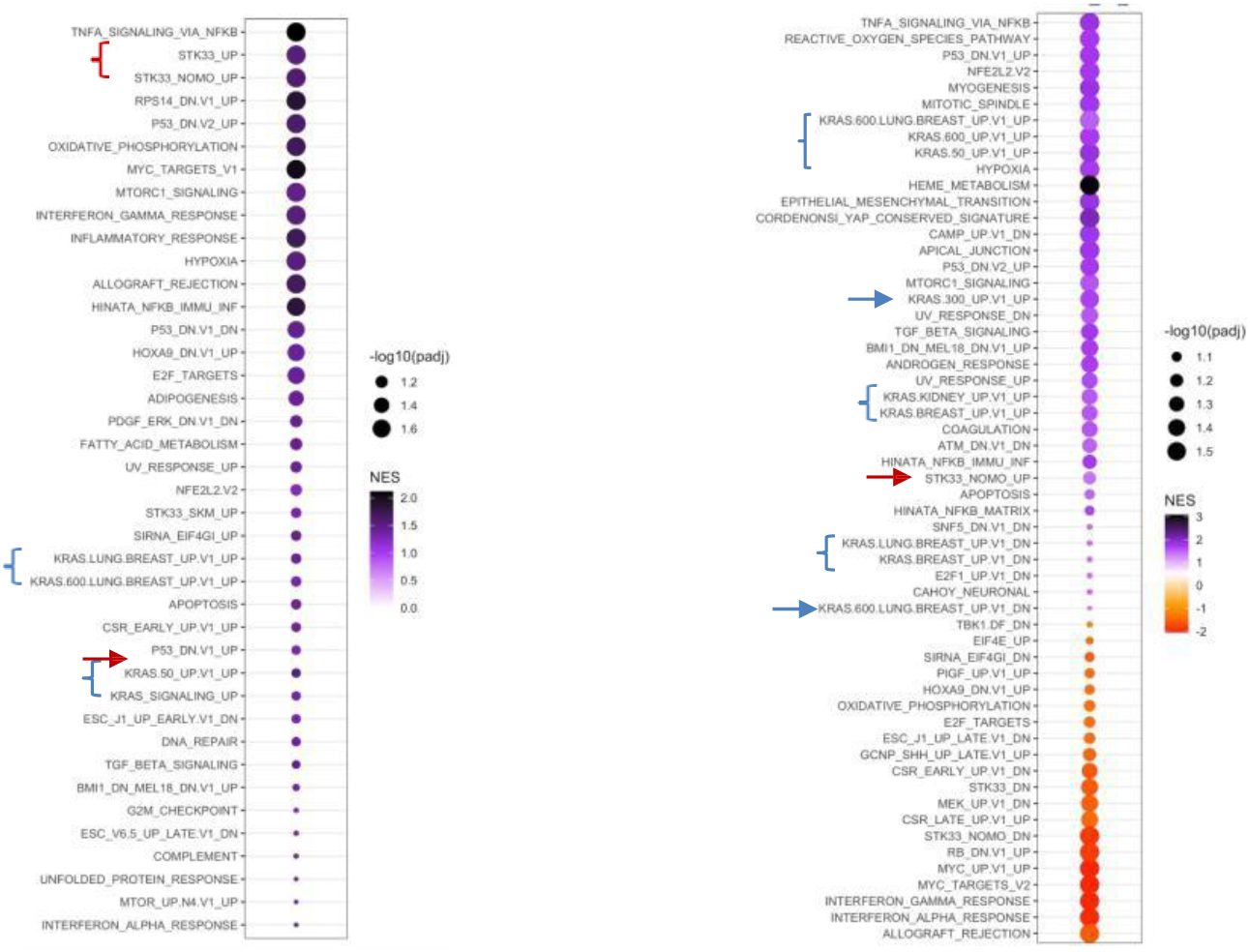
Enumeration of CTC subpopulations isolated from patients with mNSCLC. (**A**) <u>Size-based CTC enrichment schema from chemo-naive mNSCLC patients using ISET</u> <u>filters (Greek cohort, n=35)</u>. CTCs were fixed and immunostained for TfR, cytokeratin (CK), CD45 and DAPI. Following imaging CTCs were categorized as: TfR^+^/CK^-^, CK^+^/TfR^-^ and TfR^+^/CK^+^ (double positive) nucleated cells. (**B**) CTC enumeration from each subpopulation is shown as dot plot. Mann Whitney statistical test was used. ****p<0.0001. Note: patient samples with 0 CTCs are not displayed. (**C**) Kaplan Meier curve of overall survival for patients with mNSCLC dichotomized using TfR+ CTC median value (6 CTCs/sample) as the cut off. Log Rank mantel Cox statistical test was used, p=0.048. (**D**) Workflow for CTC enrichment by negative CD45^+^-cell depletion from treatment-naïve <u>mNSCLC patients (WCM cohort, n=17).</u> Enriched cells were labeled live for the cell surface proteins TfR and CD45 (leucocyte marker) and single TfR^+^-CTCs were isolated using the microscopy-based CellCelector micromanipulator. Pools of TfR^+^-CTCs, non-selected CTCs (ns-CTCs) and circulating leukocytes (PBMCs) from each patient were subjected to RNA sequencing. (**E**) Unsupervised cluster dendrogram shows TfR^+^-CTCs are transcriptionally distinct from ns-CTC population or PBMCs. (**F and G**) GSEA dot plots show pathways enriched in TfR^+^-CTCs *versus* ns-CTCs (**F**) or *versus* PBMCs (**G**). GSEA threshold was FDR≤0.05 and FC≥2. Size and color of dots represent adjusted p-value ranking and normalized enrichment score, respectively. Hallmark and Oncogenic pathways enriched in TfR^+^-CTCs are shown in purple and pathways enriched in PBMCs are shown in orange (**G**). Note that no enriched pathway was identified in the ns-CTC population (**F**). Blue symbols, KRAS oncogenic pathways; Red symbols, STK33 pathways

Additionally, we isolated CTCs from a second independent cohort of patients with mNSCLC before first-line treatment initiation (n=17, WCM cohort). In this cohort, CTCs were enriched by negative CD45 depletion using the same protocol as in patients with early-stage disease and blood was processed within 24 hr of acquisition. All 17 patients (100%) had TfR^+^-CTCs (range: 3-6782; median:127; mean: 1110) (**Supplementary Figure 5B**). The higher TfR^+^-CTC counts in the WCM metastatic cohort as compared to the Greek cohort is likely due to the different cutoffs for CTC enumeration (CTC counts in the Greek cohort were capped to 100; no cap used at WCM cohort) and possibly the different enrichment protocols. To confirm that the TfR plasma membrane labeling in the WCM cohort reflected functional TfR receptor, we incubated live CTC with fluorescently-labeled transferrin, which is the cognate ligand of TfR. Our results showed that transferrin was readily taken up by TfR^+^-CTCs and internalized, consistent with the function of TfR in transferrin-bound iron uptake (**Supplementary Figure 6**). These data further suggested that TfR captures viable, intact CTCs.

To determine the molecular features of TfR^+^-CTCs, we performed RNA-Sequencing (RNA-Seq) of up to 100 single TfR^+^-CTCs isolated from each of the 17 mNSCLC patients (WCM cohort) (**Figure 4D**). Matching PBMCs and the enriched CTC pool, after CD45 depletion and before TfR^+^ selection (non-selected CTCs), were also sequenced (**Figure 5A**). Differential gene expression analysis (DGE) and unsupervised hierarchical clustering revealed that TfR^+^-CTCs had a distinct transcriptomic profile compared with matching non-selected CTCs (ns-CTCs) or PBMCs (**Figure 4E and Supplementary Figure 7A**). Furthermore, gene set enrichment analysis (GSEA) identified several key pathways significantly enriched in TfR^+^-CTCs, such as K-Ras oncogenic signaling, p53 loss, TNFα and MTORC signaling (**Figure 4F and G**). Remarkably, there was no pathway enriched in the ns-CTC compartment compared to the TfR^+^-CTCs (**Figure 4F**), suggesting that positive CTC selection by TfR^+^ identifies unique transcriptomic networks, which would otherwise not be detectable in the pool of ns-CTCs. For example, pathways involving the STK33 serine/threonine kinase, which is both required for the survival of K-RAS mutant cells as well as targetable, were among the top three most enriched pathways in the TfR^+^-CTCs vs ns-CTCs (**Figure 4F)**. These pathways were confirmed as enriched in the TfR^+^-CTCs compared to their matching PBMCs (**Figure 4G**). Interestingly, we found only 4 pathways enriched in the ns-CTC fraction compared to matching PBMCs (**Supplementary Figure 7B).** Together, these data demonstrate that TfR^+^-CTC transcriptomic analysis can provide a snapshot of the molecular landscape of mNSCLC with potentially clinically actionable information for treatment customization.

**Figure 5.**
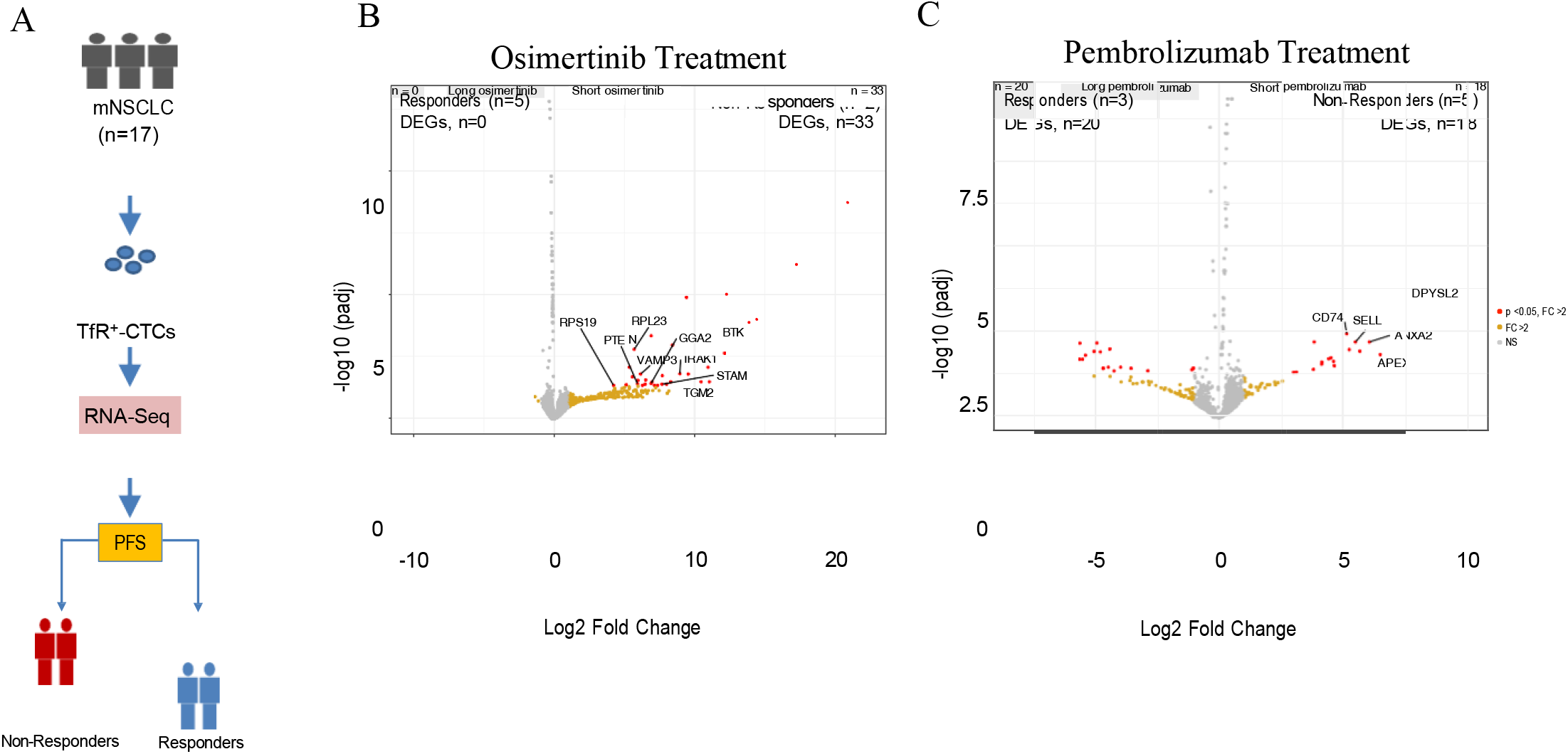
TfR^+^-CTCs transcriptomic analysis provides molecular insights into treatment response. (**A**) Study schema. RNA-sequencing of TfR^+^-CTCs isolated at baseline from patients with mNSCLC were correlated with response to treatment, using the median PFS (mPFS) from each drug’s registration trial (Osimertinib, mPFS=19 months; Pembrolizumab, mPFS=9 months) as a guide for response classification. (**B**) Of the 7 Osimertinib-treated patients we identified two subgroups of Non-Responders (n=2) with PFS of 22 days and 11 months, and Responders (n=5) with PFS of 25-36 months. (**C**) Of the 8 total patients receiving pembrolizumab-based treatment, we identified Non-Responders (n=3) with PFS of 1-9 months and Responders (n=3) with PFS of 15-34 months.

To determine whether transcriptomic analysis of TfR^+^-CTCs can provide molecular insights into treatment response, we grouped patients from the WCM cohort into those receiving targeted therapy with the EGFR tyrosine kinase inhibitor (TKI) osimertinib (n=7) and those treated with the immune-checkpoint inhibitor pembrolizumab +/- chemotherapy (n=8). Using the median historic value of progression-free survival (mPFS) for each treatment as a guide for patient response (*33, 34*), we subgrouped patients as responders and non-responders with PFS higher or lower than the historic median, respectively (**Figure 5A**). With regards to osimertinib-treated patients, TfR^+^-CTC transcriptomics identified 33 uniquely expressed genes in the non-responders (n=2) versus 0 in responders (n=5). Interestingly, 10/33 genes have been previously associated with EGFR biology and resistance to EGFR inhibitors, consistent with the patients’ quick clinical progression (**Figure 5B**). For example, BTK (Bruton tyrosine kinase) was significantly enriched in non-responders at baseline. Preclinical studies in NSCLC have shown that BTK expression confers resistance to EGFR TKIs. The increased gene expression for BTK has treatment implications because, this resistance to EGFR TKIs is readily reversible by the BTKi Acalabrutinib (*35*). Similarly, IRAK1 was significantly enriched in non-responders and was reported to confer EGFR TKi resistance *in vitro* (*36*). In contrast, transcriptomic analysis of ns-CTCs did not identify differentially expressed genes according to treatment response (**Supplementary Figure 8**). In the pembrolizumab-treated cohort (n=8) comparison of the TfR^+^-CTC transcriptomic profiles between non-responders (n=5) and responders (n=3) identified a total of 18 genes significantly enriched in non-responders. Notably, some of these genes, such as DPSYL2, ANXA2, have been directly implicated in tumor immune-cell infiltration in mNSCLC (*37, 38*). In addition, CD74 and APEX1, both known to regulate PD-L1 expression and help tumors escape immune therapy, were also significantly enriched in patients not responding to pembrolizumab treatment. (*39, 40*) (**Figure 5C**). Interestingly, small molecule inhibitors of CD74 were shown to overcome resistance to checkpoint inhibitors, such as anti-CTLA4 and anti-PD-L1 monoclonal antibodies, *in vitro* (*41*).

Taken together these data demonstrate that TfR is a reliable biomarker for CTC identification in early-stage and metastatic NSCLC. In addition, TfR+-based selection isolates viable live CTCs whose molecular characterization, prior to therapy initiation, can inform treatment selection as well as identify new actionable mechanisms of clinical drug resistance.

### TfR CTCs in pancreatic cancer

In patients with pancreatic ductal adenocarcinoma (PDAC), EpCAM-based CTC enrichment (CellSearch^®^) has identified CTCs in ∼11% to 44% of patients, albeit at low numbers (*42, 43*). However, EpCAM-based CTC enumeration did not correlate with clinical outcomes, therefore, limiting their clinical use. To test TfR’s performance in CTC identification in PDAC, we analyzed a cohort of 34 patients with advanced PDAC, and enumerated CTCs based on expression of TfR or EpCAM in the same blood sample. We found that TfR^+^ and EpCAM^+^ CTCs were detected in 100% and 94% of patients, respectively. However, TfR^+^-CTCs were the predominant CTC subpopulation (mean: 506; median: 148; range: 2-4182) compared to EpCAM^+^ (mean: 171; median: 68; range: 0-1552) (p<0.01) (**Figure 6A**). Double-positive (TfR^+^/EpCAM^+^) CTCs were also identified at low numbers (mean: 80; median: 7; range: 0-1343) (**Figure 6A and Supplementary Table 5**). Direct comparison of matching TfR^+^ *vs* EpCAM^+^-CTCs obtained from the same blood draw revealed that TfR^+^-CTCs outnumbered EpCAM^+^-CTCs for the majority of patients (70%), similar to the results in 6A (**Figure 6B and Supplementary Table 5**). Longitudinal sampling of CTCs obtained 3 months apart from patients treated with standard of care (FOLFIRINOX) (n=3), revealed that only TfR^+^-CTC counts tracked with disease severity. Specifically, we observed an increase in CTC counts upon progression, decrease upon clinical response and no change in stable disease (**Figure 6C**). By sharp contrast, enumeration of EpCAM^+^-CTC showed no correlation, suggesting that TfR^+^-CTCs are more clinically informative in patients with PDAC.

**Figure 6.**
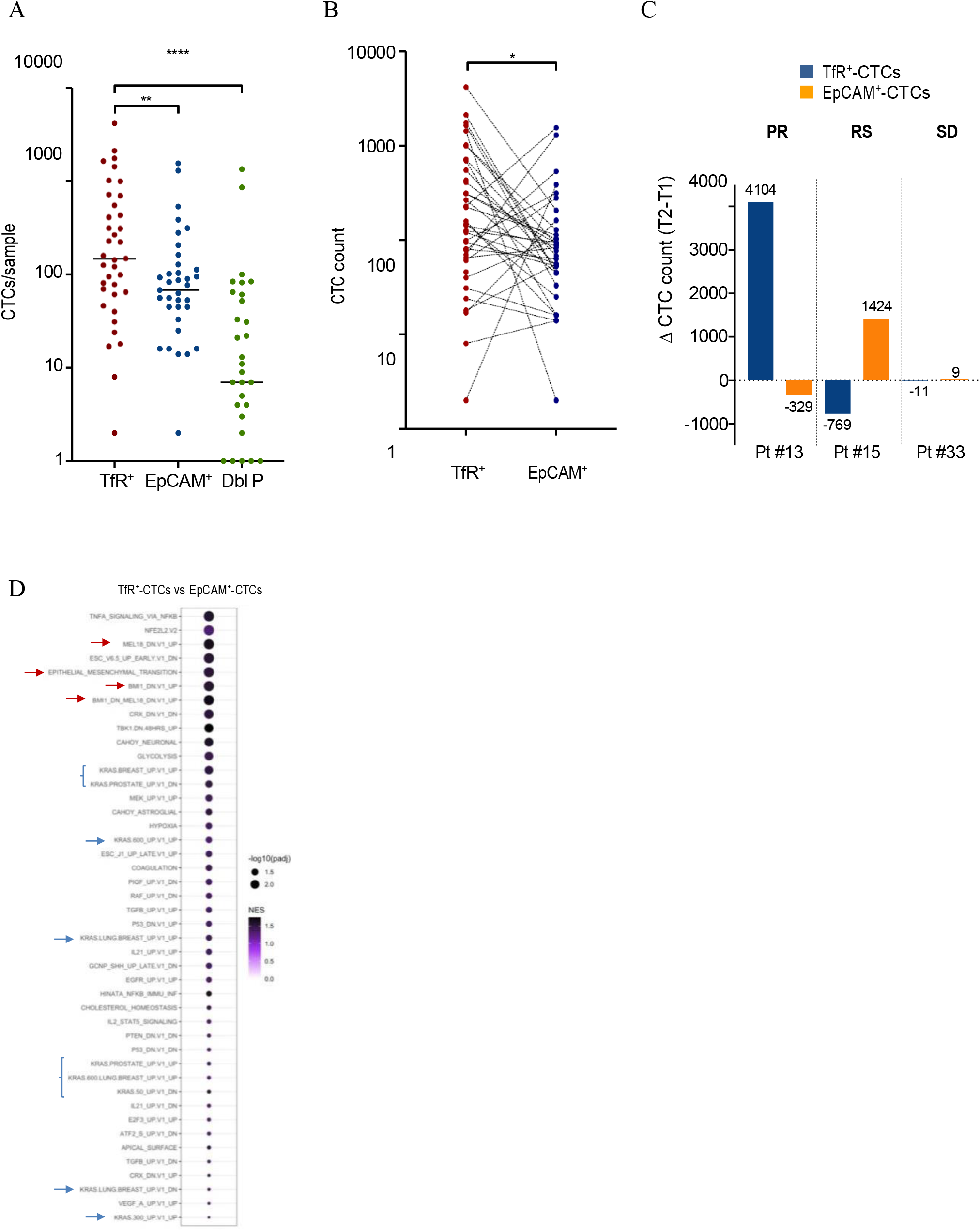
Enumeration of different immunophenotypic CTC subpopulations isolated from patients with pancreatic adenocarcinoma (PDAC). Peripheral blood was collected from patients with PDAC (n=34) and CTCs were enriched by negative depletion of CD45^+^-cells (leukocytes). CTCs were then labeled live for TfR, EpCAM and CD45 and enumerated using the Cellcelector microscopy-based micromanipulator. CTCs were identified based on size and shape (bright field); in addition to using the standard criteria of aCD45^–^ nucleated cells. CTCs were further categorized based on their respective immunophenotypes as: TfR^+^/EpCAM^-^ (TfR^+^, hereafter), EpCAM^+^/TfR^-^ (EpCAM^+^, hereafter), or double positive TfR^+^/EpCAM^+^cells. (**A**) Dot plot shows CTC counts per patient in each category. Mann Whitney statistical test was used **= p<0.01 ****= p<0.0001. (**B**) Matching TfR^+^ and EpCAM^+^ CTC counts from each patient are displayed as dot plot. Lines connect TfR^+^- and EpCAM^+^-CTCs, enumerated within each patient sample. Wilcoxon statistical test was used, *= p<0.05. (**C**) Changes in TfR^+^ and EpCAM^+^ CTC counts in three patients treated with FOLFIRINOX (standard regimen for PDAC) where blood was collected at two timepoints, T1 and 3 months later T2. The change (Δ) in CTC counts for each patient was calculated as T2-T1 counts. Y axis displays the CTC count Δ for each CTC category (TfR^+^ vs EpCAM^+^) and the exact values are displayed for each patient and respective category. Positive values indicate increase in CTC counts at T2 while negative values indicate decrease in CTC counts at T2. The difference in CTC counts, also displayed in the graph) was then correlated with disease status at T2; PR= progression; RS= response; SD= stable disease. (**D**) Transcriptomic analysis in TfR^+^-*versus* EpCAM^+^-CTCs. CTCs were enriched labeled live as in A, from a subset of the PDAC patients (n=12/34). Single CTCs were isolated based on TfR^+^ or EpCAM^+^ expression using the CellCelector micromanipulator. Pools of TfR^+^-CTCs and EpCAM^+^-CTCs were then subjected to RNA sequencing. GSEA dot plot shows pathways enriched in TfR^+^-CTCs *versus* EpCAM^+^-CTCs. Results are ranked based on adjusted p-value and color-coded based on normalized enrichment score. Hallmark and Oncogenic pathways enriched in TfR^+^-CTCs are shown in purple. Note that there was no enriched pathway in EpCAM^+^-CTCs.

Transcriptomic analysis of TfR^+^ *vs* EpCAM^+^ -CTCs (isolated within the same blood draw, n=12) identified 248 genes significantly enriched in TfR^+^-CTCs compared to 44 unique genes in EpCAM^+^-CTCs (**Supplementary Figure 9**). GSEA analysis further revealed that TfR^+^-CTCs, compared to EpCAM^+^-CTCs, were enriched in oncogenic pathways known to drive PDAC progression, such as K-Ras, Raf and MEK pathways, (**Figure 6D**). It is interesting to note that we did not detect any pathway enriched in EpCAM^+^-CTCs suggesting that no oncogenic network in particular was represented in this CTC subpopulation. Additional TfR^+^-specific pathways were associated with inflammation (TNF-α) and an EMT phenotype (epithelial_mesecnhymal_transition; MEL18_DN), consistent with the clinical manifestations of PDAC.

## Discussion

Circulating tumor cells have emerged as a reliable source of tumor tissue readily accessible from the peripheral blood of patients with solid tumors to perform cell-based assays and dissect the molecular profile of the disease. However, CTC clinical utility is currently limited by the rarity of this blood cell population in the blood stream and the technical limitations associated with their isolation and identification. Currently, the FDA-cleared technologies for CTC isolation (CellSearch^®^, Menarini) and molecular analysis (ADNATest) rely on the expression of EpCAM as a cell surface antigen for CTC isolation from the peripheral blood of cancer patients. However, the clinical use of these assays is limited to few cancer types, and it is restricted to a prognostic role. Antigen-agnostic CTC identification methodologies such as Epic Sciences are commercially available. However, the fixation requirements of this platforms restrict its clinical applications to CTC morphometric characterization and a narrow genomic profiling, thus limiting the possibility to perform more complex molecular CTC phenotyping, such as untargeted transcriptomic characterization.

Other markers have recently been tested as alternative to EpCAM for CTC identification and isolation (*i.e.*, folate receptor or VAR2CSA) (*56, 57*). Despite the fact that these cell-surface proteins can be successfully used to capture CTCs from patients with solid tumors, the clinical relevance of their enumeration as a prognostic tool and of their molecular characterization has yet to be fully explored.

In this study, we investigated the role of TfR as a cell surface biomarker for the identification of CTCs in different tumor types. TfR functions as the mediator of intracellular iron intake; it is ubiquitously expressed in normal tissue, with increased expression in metabolically active tissues characterized by high iron requirements (*i.e*., intestinal epithelium, placental trophoblast) (*14*). TfR overexpression has been repeatedly reported in cancer cells as compared to normal tissue, likely reflecting the increased metabolic and iron requirements of cancer cells (*44, 45*). Importantly, several oncogenic growth and transcription factors, such as c-Myc (*46*), HIF-1 α (*47*) and Src (*48*), promote TfR over-expression in cancer cells to support their high proliferation rate.

The selective overexpression of TfR in cancer cells, the clinical association of TfR with advanced disease stage and worse prognosis in solid tumors, and its plasma membrane localization (*49, 50*) have made TfR an attractive target as a diagnostic and therapeutic resource (*51–53*). These characteristics also make TfR also an ideal candidate for the identification and isolation of CTCs.

Our results indicate that TfR identifies a larger number of CTCs in the peripheral blood of patients with different types of solid tumors, as compared to EpCAM. TfR^+^-CTC counts were significantly higher than standard epithelial markers in cancers where EpCAM-based CTC counts have clinical relevance, such as prostate cancer, as well as in tumor types where EpCAM-based enumeration has marginal or no clinical impact, such as pancreatic cancer and NSCLC. Importantly, we showed that enumeration of TfR^+^-CTCs, but not EpCAM^+^-CTCs, is associated with disease progression and overall survival, in prostate cancer and NSCLC, respectively. These results, together with the fact that TfR expression is not affected by epithelial-to-mesenchymal transition, support the use of TfR as a marker for positive selection of CTCs across different tumor types.

Interestingly, TfR was able to identify CTCs in patients with early-stage NSCLC, a clinical setting where liquid biopsies are much needed to improve patient stratification. Previous studies detected CTCs in the peripheral blood of patients with early-stage NSCLC using either EpCAM-based or size-based CTC isolation platforms; however, the CTC detection rate reported in these studies was lower (22-50%) than what we report here using TfR as a marker for CTC isolation (100%) (*54, 55*). Taken together, these results encourage the future investigation of TfR-based CTC isolation in early-stage NSCLC.

Our study not only showed that TfR^+^-CTCs are significantly more abundant than EPCAM^+^-CTCs, but also demonstrated that they possess a unique molecular transcriptomic profile, which can inform potential mechanisms of resistance to standard treatments (*4, 22, 32*). The FDA-cleared EpCAM-based Adna-test AR-V7 detect platform is commercially available to detect the presence of AR-V7 in EpCAM-positive CTCs, and predicts response to AR signaling inhibitors in prostate cancer (*4*). Our data here show that TfR^+^-CTCs are enriched in the expression of the two clinically relevant AR variants, V7 and v567, as compared to EpCAM^+^-CTCs, suggesting that TfR identifies a clinically relevant population of CTCs whose molecular interrogation could help clinicians select the optimal treatment in patients with prostate cancer.

The transcriptomic profiling of TfR^+^-CTCs shed light on their biology and clinical role in disease progression and response to treatment. TfR^+^-CTC transcriptomics were enriched in K-Ras related pathways in both NSCLC and pancreatic cancer. K-Ras pathway activating mutations are the most common tumor molecular alterations in lung cancer (*58*) and K-Ras inhibitors are predominant therapeutic strategies currently emerging for lung cancer treatment (*59*). The enrichment of several K-Ras associated pathways in TfR^+^-CTC, together with the correlation between TfR^+^-CTC enumeration and survival in mNSCLC, underline the biological and clinical relevance of TfR^+^-CTC in progression and clinical outcome of this disease.

Notably, TNF-α signaling was the most enriched pathway in TfR^+^-CTCs in both NSCLC and PDAC. Previous associations of high TNF-α expression with metastatic spread and treatment resistance in both tumor types (*60–62*) underline the clinical relevance of TfR^+^-CTCs in tumor progression, and support future investigation to clarify the role of this CTC subpopulation in tumor dissemination. Interestingly, the Nuclear Factor Erythroid-derived 2-like2 (NFE2L2) related network was one of the most highly enriched pathways in TfR^+^-CTCs as compared to ns-CTCs or EPCAM^+^-CTCs in lung cancer and in PDAC, respectively. Preclinical studies have shown that pancreatic cancer initiation is supported by K-Ras driven NFLE2L2 activation, which is required to support PDAC cells proliferation, and that pharmacological inhibition of AKT can suppress NFE2L2-driven tumor proliferation (*63*). Notably, NFE2L2 has also been shown to play a crucial role in lung cancer progression and metastasis (*64*). It was also interesting to note that TfR^+^-CTCs were enriched in pathways associated with epithelial-to-mesenchymal transition (EMT) and stemness, such as the BMI1/MEL18 axis pathways (*65, 66*); these clinical transcriptomic features are in line with our preclinical data showing that TfR expression is not affected by EMT induction and support the hypothesis that TfR can capture CTCs across the entire EMT spectrum.

To our surprise, we did not find any pathways selectively upregulated in non-selected CTCs (ns-CTCs) in NSCLC compared to TfR^+^-CTCs, and only one K-Ras-related pathway was enriched in ns-CTCs when the ns-CTCs were compared to PBMCs in this disease. These data suggest that TfR^+^-CTCs transcriptomic profile reveals clinically relevant, potentially targetable molecular pathways, which would otherwise be missed and deemed not relevant if we would limit our molecular analysis to non-selected CTCs. Similarly, no pathways were selectively enriched in the purely epithelial EpCAM^+^-CTCs in PDAC, further corroborating the clinical relevance of TfR^+^-CTCs detection and molecular characterization in solid tumors.

Our transcriptomic analysis showed that molecular profiling of TfR^+^-CTCs can be informative of treatment response to standard therapies in NSCLC (*i.e.*, EGFR-TKIs and immune checkpoint inhibitors) and can help identify genes potentially implicated in intrinsic clinical resistance. In our cohort of seven NSCLC patients who received osimertinib as first-line of treatment, the transcriptomic analysis of TfR^+^-CTCs at baseline showed upregulation of BTK, which confers resistance to EGFR TKIs, in the two non-responders as compared to the five responders. Our findings confirm, for the first time clinically, the role of BTK expression in intrinsic clinical osimertinib resistance and offer an alternative pathway for treatment with the clinically approved available BTKis. Previous clinical studies showed BTK-TKis have limited clinical activity for the treatment of relapsed NSCLC (NCT02403271, NCT02448303). However, these studies mostly focused on the combination of BTK-TKi with immune checkpoint inhibitors, failing to address the potential benefit of BTK-TKI/EGFR-TKIs combination regimens.

## Data Availability

All data produced in the present study are available upon reasonable request to the authors

## Acknowledgments

**Funding**

Department of Defense grant W81XWH-18-PCRP-IDA (to P.G.)

National Cancer Institute of the National Institutes of Health, R01 CA179100 (to P.G.)

National Cancer Institute of the National Institutes of Health, R01 CA137020 (to P.G.)

National Cancer Institute of the National Institutes of Health, R21 CA216800 (to P.G.)

National Cancer Institute of the National Institutes of Health, T32CA062948 (to G.G)

National Cancer Institute of the National Institutes of Health, T32CA203702 (to J.Z.)

## Author contributions

Conceptualization: PG, TEM

Funding acquisition: PG

Investigation: GG, AK, AG, JZ, BZ, RB

Resources: ST, AO, GK, DN, NA, AS

Writing-Original Draft: GG, PG

Writing-review & editing: GG, PG, NA, ST, DN, AS, AO, TEM

Supervision: PG, NA, ST, DN, AS, AO, TEM

### Data and material availability

All data are available in the main text or the supplementary materials

## Supplementary Materials

